# Diagnostic Performance of Rapid Antigen Testing for SARS-CoV-2: The COVid-19 AntiGen (COVAG) Extension study

**DOI:** 10.1101/2023.11.29.23299183

**Authors:** Christoph Wertenauer, Alexander Dressel, Eberhard Wieland, Hans-Jörg Wertenauer, Helmine Braitmaier, Anna Straub, Nicholas Lützner, Winfried März

## Abstract

**Background:** Rapid antigen tests (RATs) for SARS-CoV-2 have been used to combat the still ongoing Covid-19 pandemic. This study is the extension of the COVAG study originally performed from February 1 to March 31, 2021. We compared two RATs, the Panbio COVID-19 Ag Rapid Test (Abbott) and the SD Biosensor Q SARS-CoV-2 Rapid Antigen Test (Roche), against RT-PCR on the foil of new variants.

**Methods:** We included 888 all-comers at a diagnostic center between October 20, 2021, and March 18, 2022. RT-PCR-positive samples with a Ct value ≤ 32 were examined for SARS-CoV-2 variants.

**Findings:** The sensitivity of the Abbott-RAT and Roche-RAT were 65% and 67%, respectively. For both RATs, lower Ct values were significantly correlated with higher sensitivity. For samples with Ct values ≤ 25, the sensitivities of the Roche-RAT and of the Abbott-RAT were 96% and 95%, for Ct values 25-30 both were 19%, and for Ct values ≥ 30 they were 6% and 2%, respectively. The RATs had substantially higher sensitivities in symptomatic than asymptomatic participants (76, 77%, vs. 29, 31%, for Abbott-RAT, Roche-RAT, respectively) and in participants referred to testing by their primary care physician (84%, 85%) compared to participants who sought testing due to referral by the health department (55%, 58%) or a warning by the Corona-Warn-App (49%, 49%). In persons with self-reported previous Covid-19 sensitivities were markedly lower than in patients without previous Covid-19: 27% vs. 75% for Roche-RAT and 27% vs. 73% for Abbott-RAT. Depending on the vaccination status, the sensitivity of the RATs is 67.6%, 61.5% and 70.6% for non-vaccinated, vaccinated and boostered participants, respectively. For the considered subpopulation of 888 participants, we find no significant correlation between vaccination status and sensitivity.

The Omicron variant was detected with a sensitivity of 94% and 92%, the delta variant with a sensitivity of 80% and 80% for Abbott-RAT and Roche-RAT, respectively. This difference is attributable to the lower Ct values of the Omicron samples compared to the Delta samples. When adjusted for the Ct value, a multivariate logistic regression did not show a significant difference between Omicron and Delta. In terms of sensitivity, we found no significant difference between the wild-type and the Omicron and Delta variants, but a significantly lower sensitivity to the alpha variant compared to the other variants.

For a Ct value ≤ 25 the sensitivities were 95.2% and 96.0% for the Abbott-RAT and the Roche-RAT, respectively (Table 4). For a Ct value of 25-30 both RATs had a sensitivity of 18.8%. For a Ct value of 30-32, the sensitivities were 0.0% and 7.1% respectively, for Ct values ≥32 the sensitivities were 3.0% and 6.0% for Abbott-RAT and Roche-RAT, respectively.

The specificities were >99% overall.

Interpretation: The sensitivity of the RATs for asymptomatic carriers is unsatisfactory questioning their use for screening. When used in symptomatic patients or when requested by a primary care physician the sensitivities were higher. Our study does not suggest that the vaccination status influences the sensitivity of RATs.

## 1. Introduction

Severe acute respiratory syndrome Coronavirus type 2 (SARS-CoV-2) is the causative agent of Coronavirus disease 19 (Covid-19). Covid-19 emerged in late 2019, quickly spread around the world and was declared a global pandemic on March 11, 2021, by the World Health Organization (WHO).^1^ Since its emergence multiple SARS-CoV-2 variants developed which mostly were characterized by mutations in the Spike protein but also within the Nucleocapsid protein.^2–5^ Variants showing a decrease in the effectiveness of available diagnostic tests among other criteria are termed Variants of Concern (VOC) by the WHO.^6^ To date the WHO has listed 5 VOCs, namely: B.1.1.7 (alpha), B.1.351 (beta), P.1 (gamma), B.1.617.2 (delta) and the currently prevailing B.1.1.529 (Omicron).^6^ For Omicron several sub-lineages have been identified with BA.5 being the currently dominant one in Europe.^3^

The clinical presentation of Covid-19 ranges from asymptomatic to prolonged illness requiring intensive care treatment and death.^7,8^ As SARS-CoV-2 can be transmitted by symptomatic as well as asymptomatic persons the identification of infectious carriers is crucial to contain Covid-19 by means of contact tracing and isolation of infectious patients.^8^ This requires effective testing and an early diagnosis of Covid-19. Detection of acute SARS-CoV-2 infection can be achieved by direct testing including nucleic acid amplification tests (NAATs) or through rapid antigen tests (RATs). NAATs identify viral RNA in specimens from the respiratory tract while RATs recognize viral proteins, mostly the Nucleocapsid protein.^9^ To date NAAT-based assays such as reverse transcription-polymerase chain reaction (RT-PCR) are the gold standard in detecting acute SARS-CoV-2 infection. RATs are widely employed as well as they can be conducted at the point of care, provide fast results within 15-30 minutes, and can be used for self-testing. Positive RAT results need to be verified by RT-PCR testing.^10^ Indirect tests including assays detecting antibodies against the Spike- or the Nucleocapsid protein are not useful in the diagnosis of acute infection as they only become positive after 3 days and more or may be already positive from an earlier infection (Nucleocapsid- and Spike antibodies) or vaccination (Spike-Antibodies).^11,12^

This study is the extension of the COVAG study originally performed from February 1, 2021, to March 31, 2021. During the first data collection period we saw that the alpha variant decreased the effectiveness of the RATs compared to the wild-type.^13^ As new SARS-CoV-2 variants emerged afterwards, the COVAG study was continued to comprehensively examine two of the most sensitive RATs in a real-world, prospective, head-to-head study, placing specific emphasis on clinical characteristics and the presence of SARS-CoV-2 variant genotypes.^9^

## 2. Methods

### 2.1. Setting and Participants

This prospective study was conducted at the Corona Test Centre Cannstatter Wasen in Stuttgart, Germany as an extension of the COVAG study.^13^ Individuals scheduled for RT-PCR testing of nasopharyngeal swabs were advised of the study orally and in writing. Participants had to be aged ≥ 18 years and capable of understanding the nature, significance, and implications of the study. Children and adolescents <18 years of age and patients obviously suffering from clinical conditions requiring emergency hospitalization were excluded. All participants provided written and informed consent. The study was approved by Ethics Committee II (Mannheim) of the University of Heidelberg (reference number 2020-417MF) and the German Institute for Drugs and Medical Devices.

We recorded demographic characteristics, reasons for testing, medical history including SARS-CoV-2 vaccination history, clinical symptoms, and vital signs (heart rate, blood pressure, body temperature, and oxygen saturation) and we stratified the reasons for testing into four major categories: participants referred by their primary care physicians, by the Health Department, participants seeking RT-PCR testing to confirm a positive antigen test and participants who received a warning in their digital contact-tracing app (Corona-Warn-App). We grouped the participants based on their Covid-19 vaccination status into non-vaccinated (0 or 1 vaccination), vaccinated (2 vaccinations), boostered (3 or more vaccinations) and with unknown vaccination status. In addition to collecting the oro- and nasopharyngeal swabs for RT-PCR testing, we collected two completely independent nasopharyngeal swab specimens to run two commercially available and widely used RATs. The swabs were collected by medically educated personnel of the test center by rotating teams with strict adherence to the instructions issued by the manufacturers. We used the Abbott Panbio^TM^ COVID-19 Ag Rapid Test (Abbott Rapid Diagnostics Jena GmbH, Jena, Germany, www.abbott.com/poct) and the Roche-SD Biosensor SARS-CoV-2 Rapid Antigen Test (identical to SD BIOSENSOR Standard Q COVID-19 Ag, www.sdbiosensor.com; Roche Diagnostics; Mannheim, Germany, www.roche.com). We chose those two tests in continuation of our first data collection period and because they were among the most sensitive tests according to a Cochrane analysis.^13,14^

Hereafter, we refer to the tests as Abbott-RAT and Roche-RAT, respectively. We randomly assigned the participants to three sampling groups according to the sequence of collecting the nasopharyngeal swabs (group 1: RT-PCR, RAT-Roche, RAT-Abbott; group 2: RAT-Roche, RAT-Abbott, RT-PCR; and group 3: RAT-Abbott, RT-PCR, RAT-Roche) to reduce bias based on the order of test performance.

### 2.2. Analytical procedures

Both the Abbott-RAT and the Roche-RAT were carried out by medically educated staff according to the manufacturers’ instructions on-site at the Corona Test Centre, immediately after sampling the nasopharyngeal swabs. The nasopharyngeal swabs for real-time RT-PCR (rRT-PCR) testing were placed in 2 ml of a phosphate-buffered saline solution (ISOTON™ II Diluent, Becton Dickinson, Galway, Ireland) and delivered to the SYNLAB Medical Care Centre Leinfelden-Echterdingen. This ensured that the performers of the RATs were unaware of the RT-PCR-results.

SARS-CoV-2 RNA was extracted from the nasopharyngeal swab samples and purified using the PurePrep Pathogens kit and a PurePrep 96 instrument (Molgen, Veenendaal, the Netherlands) within 6 h after sampling to limit degradation. The integrity of the RNA was monitored by co-amplification of a control RNA included in the solution for the lysis of the swabs. In cases in which neither SARS-CoV-2 RNA nor the control RNA were detected, the RNA preparation was repeated. The rRT-PCR assay was performed using either the RIDA®GENE SARS-CoV-2 test kit (R-Biopharm, Darmstadt, Germany) or the Allplex SARS-CoV2 (Seegene, Seoul, Korea) or the Virella SARS-CoV-2 seqc (Gerbion, Kornwestheim, Germany) on the CFX96 Touch Real-Time PCR detection device (Bio-Rad, Feldkirchen, Germany) or the CFX-96 IVD Real-Time PCR detection device (Bio-Rad, Feldkirchen, Germany) according to the manufacturers’ instructions. The RIDA®GENE SARS-CoV-2 test kit targets the SARS-CoV-2 envelope (E) gene, the Allplex SARS-CoV2 targets the N-gene, S-gene/RdRP and the E-gene (pan Sarbecovirus) and the Virella Seqc SARS-CoV2 targets the RdRp/S-gene and the E-gene (pan Sarbecovirus). Samples producing a cycle threshold (Ct) ≤ 35 were considered positive by RT-PCR.

We screened RT-PCR-positive samples with a Ct ≤ 32 for SARS-CoV-2 variants of concern (VOC). Until November 8^th^ 2021 this analysis was performed at SYNLAB Medical Care Center Leinfelden-Echterdingen using the Kits Seegene Allplex Variant I (Seegene, Seoul, Korea) and Virella SARS-CoV2 Mut 3 (Gerbion, Kornwestheim, Germany) according to the supplier’s instructions. Afterwards the VOC analysis was performed at SYNLAB Medical Care Center Weiden using the Novaplex SARS-CoV-2 Variants I Assay, Novaplex SARS-CoV-2 Variants IV Assay and Novaplex SARS-CoV-2 Variants VII Assay (Seegene, Seoul, Korea) according to the supplier’s instructions.

Samples were screened for B.1.617.2 (delta), B.1.617.2.1 (delta plus), B.1.1.529 / BA.1 (omicron) and BA.2 (omicron stealth). Samples with positive results for L452R and P681R and absence of K417N were assigned to the delta variant. Positive results for L452R, P681R and K417N were considered as Delta plus. Presence of N501Y, E484A and HV69/70del were considered as Omikron BA.1 and occurrence of N501Y, E484A with absence of HV69/70del as Omikron BA.2.

Additionally, 378 RT-PCR-positive samples with a Ct ≤ 30 were sequenced in April and Mai, 2022 at the SYNLAB Medical Care Center Weiden and SYNLAB Medical Care Center Mannheim using the Illumina COVIDSeq Test according to the supplier’s instructions. 98 SARS-CoV-2 amplicons were sequenced for each sample. This Whole-Genome-Sequencing (WGS) approach was performed using the NextSeq 500/550/550Dx (Illumina, San Diego, USA) and NovaSeq 6000 systems (Illumina, San Diego, USA). The obtained sequencing reads of RNA libraries, prepared using the ARTIC v3 gene assay panel (Illumina COVIDSeq Test, Illumina, San Diego, USA), were analyzed with the DRAGEN COVID Pipeline (Illumina, San Diego, USA). Third-party software applications Pangolin (version 4.0.6/UsHER Covid-Pipeline_1.1.0) and NextClade (version 1.11.0) were used for lineage and clade determination. Only samples with ≥95% genome coverage (% of non-N bases) were statistically processed.

### 2.3. Statistical Analysis

Continuous data are presented as means, standard deviations (SD), medians, and 25th and 75th percentiles. Categorical data are presented as absolute numbers and percentages (Table 1).

In our analysis, the performance indicators for the two RATs in relation to RT-PCR (chosen as the gold standard for having COVID-19) are given by sensitivity, specificity, positive predictive value (PPV), negative predictive value (NPV), and diagnostic efficacy (number of correct test results divided by the total number of test results).

In Table 2, the p-values apply to two-sided testing of the null hypothesis that the difference between the Abbott-RAT and the Roche-RAT performance indicators is equal to zero. The probability densities underlying the two-sided testing are estimated by means of 5000 bootstrap iterations.

The risk of having COVID-19 according to baseline anthropometric and anamnestic characteristics was expressed in terms of crude odds ratios (ORs) and ORs adjusted for age and sex as calculated by logistic regression (Supplementary table 1).

We also visualized the sensitivities of both RDTs relative to the rRT-PCR-derived Ct values (Figure 2) and the PPVs and NPVs according to hypothetical disease prevalence rates in the range of 0-0·05 (Figure 3). To compare the PPV and NPV of the RDTs with standardized criteria on performance, we also used the following hypothetical sensitivity and specificity levels (tiers 1-3) recommended by Kost et al. (34): tier 1, 90%, 95%; tier 2, 95%, 97.5%; and tier 3, 100%, ≥99% (Figure 3).

Finally, we investigated whether the sensitivities of the two RDTs were related to the reason for testing, comorbidities, clinical symptoms, vital signs, or SARS-CoV-2 genotypes using univariate (Table 2) and multivariate logistic regression (Table 3).

The statistical tests were two-sided and P<0·05 was considered significant. The analyses were carried out using R v4.0.2 (http://www.r-project.org).

## 3. Results

### 3.1 Clinical characteristics of Participants

The extension of the COVAG study was conducted from October 20, 2021 to March 18, 2022. Figure 1 shows the data collection period and the emergence of variants framed within the course of the COVID-19 pandemic in Germany. A total of 1508 persons agreed to participate in this study. 21 persons were disregarded from further evaluation because at least one of the three tests was not available. This resulted in 1487 persons enrolled in the COVAG Extension study (Figure 2, Supplementary table 1) including 801 (53.9%) women, 685 (46.1%) men and one diverse person (0.1%). Adverse effects from performing any of the tests were not experienced.

**Figure 1:**
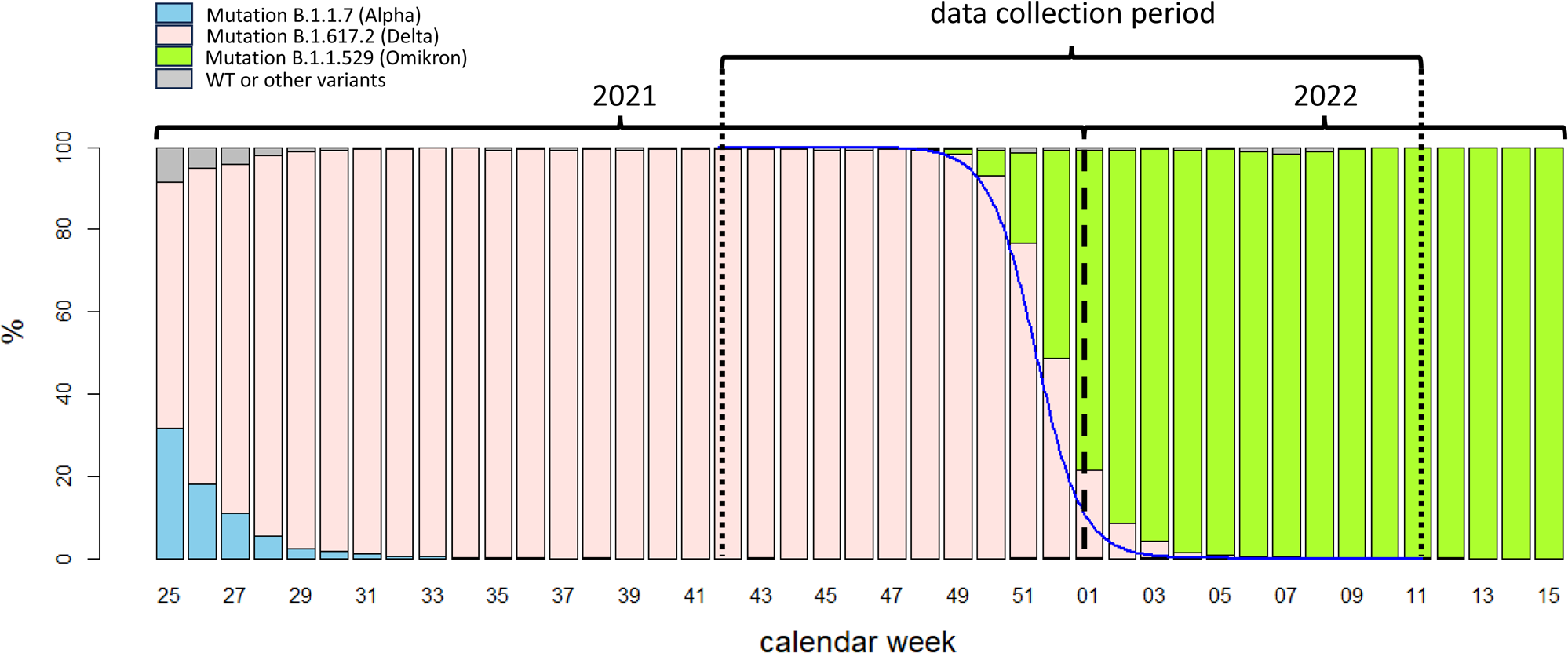
Framing of the COVAG extension study (October 20, 2021, to March 18, 2022) into the time course of the COVID-19 pandemic in Germany. *Abszissa:* calendar week within 2021 and 2022; *bars:* Germany-wide weekly proportions of variants of concern (VOC) in percent. *Blue solid line:* estimated proportion of variant B.1.617.2 (Delta) in the COVAG extension study to Germany (based on logistic regression with the categories ‘Delta’ vs. ‘Omikron’).

**Figure 2:**
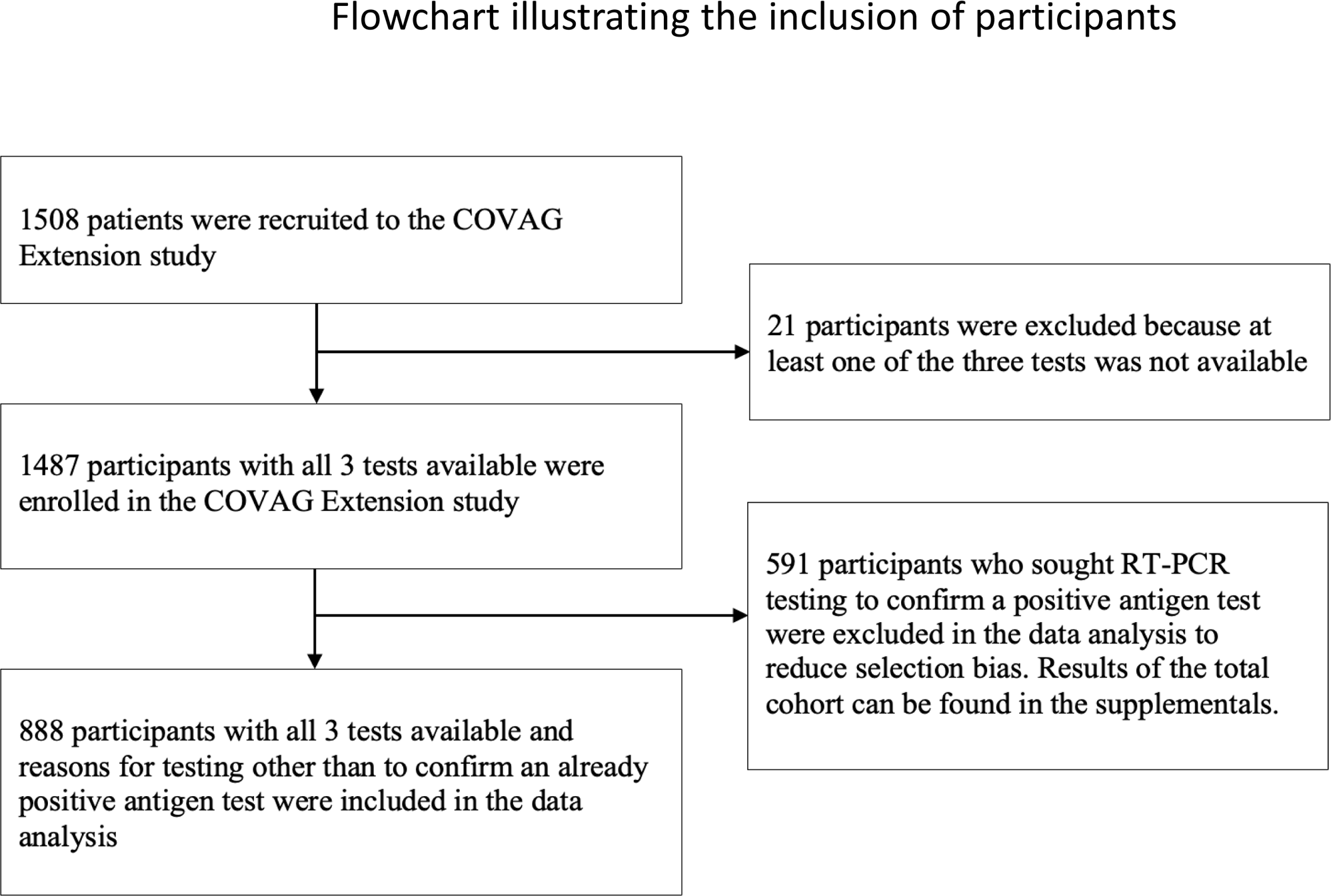
Flowchart illustrating the inclusion of participants into the COVAG Extension study and data analysis.

Within the period of data collection, self-testing with RATs and RT-PCR confirmation in the case of a positive RAT was performed very frequently in Germany which in many participants who already had a positive self-test beforehand. To reduce selection bias, we excluded these 591 (39.7%) participants from the further analyses (Figure 2). 888 participants were tested for other reasons. Those included a warning by the Corona-Warn-App in 419 (28.2%) participants, a referral from the health department in 236 (15.9%) participants, and a referral from the primary care physician in 233 (15.7%) participants. For 8 (0.5%) participants no information regarding the reason for testing was available (Table 1). The anthropometric and anamnestic characteristics of all 1487 participants can be found in Supplementary table 1. Further data analysis was performed for the 888 participants with reasons for testing other than to confirm a positive RAT.

Of 888 participants, 497 (56%) were women and 390 (43.9%) were men, one person (0.1%) is assigned neither to women nor to men. 665 (74.9%) participants self-reported having no comorbidities, while 223 (25.1%) reported having any comorbidities. The most common comorbidities were hypertension (9.5%) and dyslipoproteinemia (4.7%). Other comorbidities were low in frequency. 101 (11.4%) participants self-reported having had a previous Covid-19 infection (Table 1).

98 (11.0%) participants are non-vaccinated (0 or 1 vaccination against Covid 19), 321 (36.2%) participants are ‘vaccinated’ (2 vaccinations against Covid 19), 463 participants (52.1%) have received a booster vaccination (3 or more vaccinations against Covid 19). For six persons (0.7%), the vaccination status is unknown.

447 (50.3%) participants reported having clinical symptoms while 441 (49.7%) reported none. The most common symptoms were malaise, cough, headache, and musculoskeletal pain at frequencies of 36.6 %, 30.3%, 30.0%, and 16.3%, respectively (Table 1).

188 (21.2%) participants were tested positive for SARS-CoV-2 by RT-PCR. 126 (14.2%) were tested positive by the Abbott-RAT and 128 (14.4%) by the Roche-RAT. 125 (14.1%) samples had a Ct value ≤25, 16 (1.8%) a Ct value 25-30, 47 (5.3%) ≥ 30. 155 (17.5%) RT-PCR samples had a Ct value ≤ 32. 52 RT-PCR positive samples with a Ct value ≤ 32 could not examined be for variants. Of the remaining 103 samples, the Omicron variant was found in 41 and the Delta variant was found in 62.

### 3.2 Diagnostic performance of RATs

#### Sensitivity

The Abbott-RAT and the Roche-RAT had overall sensitivities of 65.4% (95% CI 60.7-70.2%) and 67.0% (95% CI 62.4-71.8%) respectively (Table 2). The sensitivities of both RATs were significantly associated with the Ct-value derived from RT-PCR (Figure 3A).

**Figure 3:**
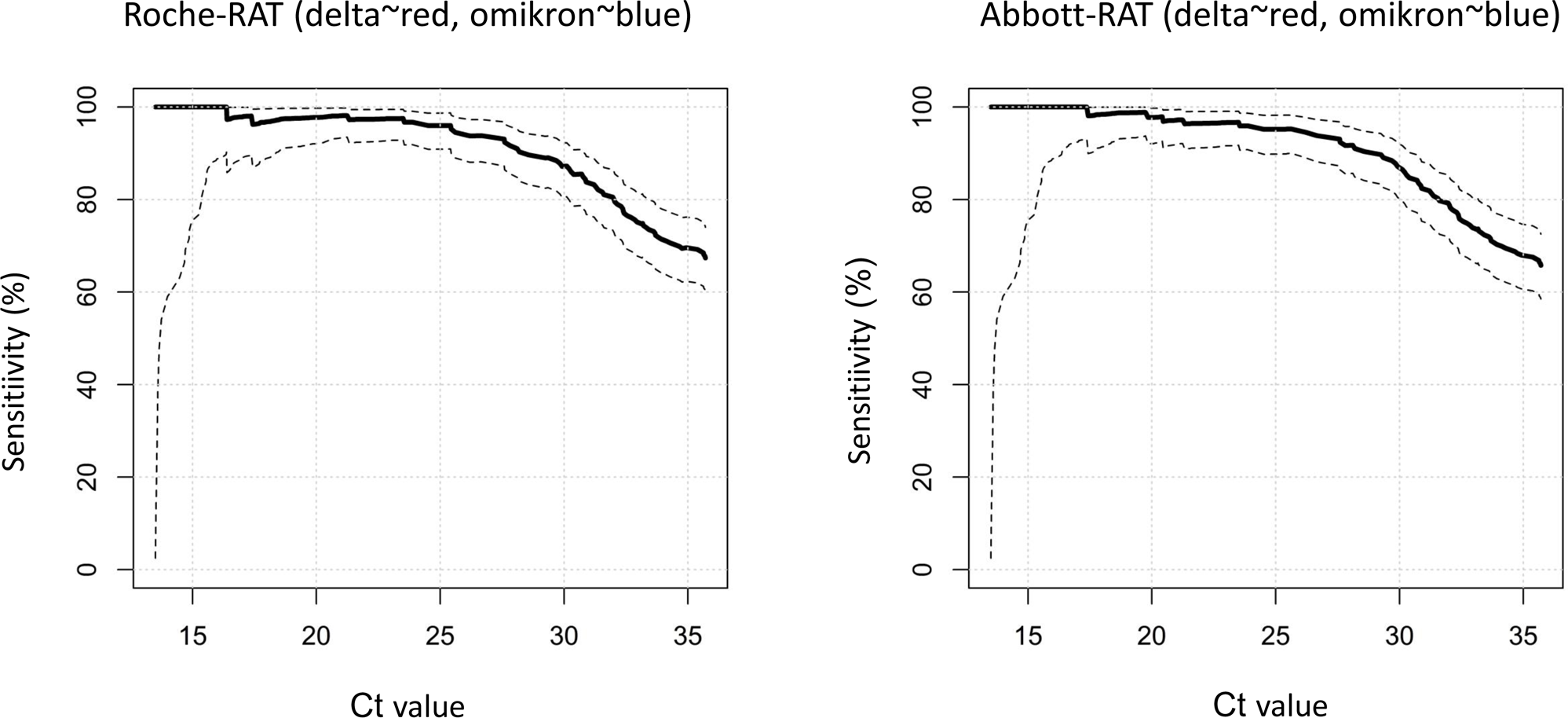
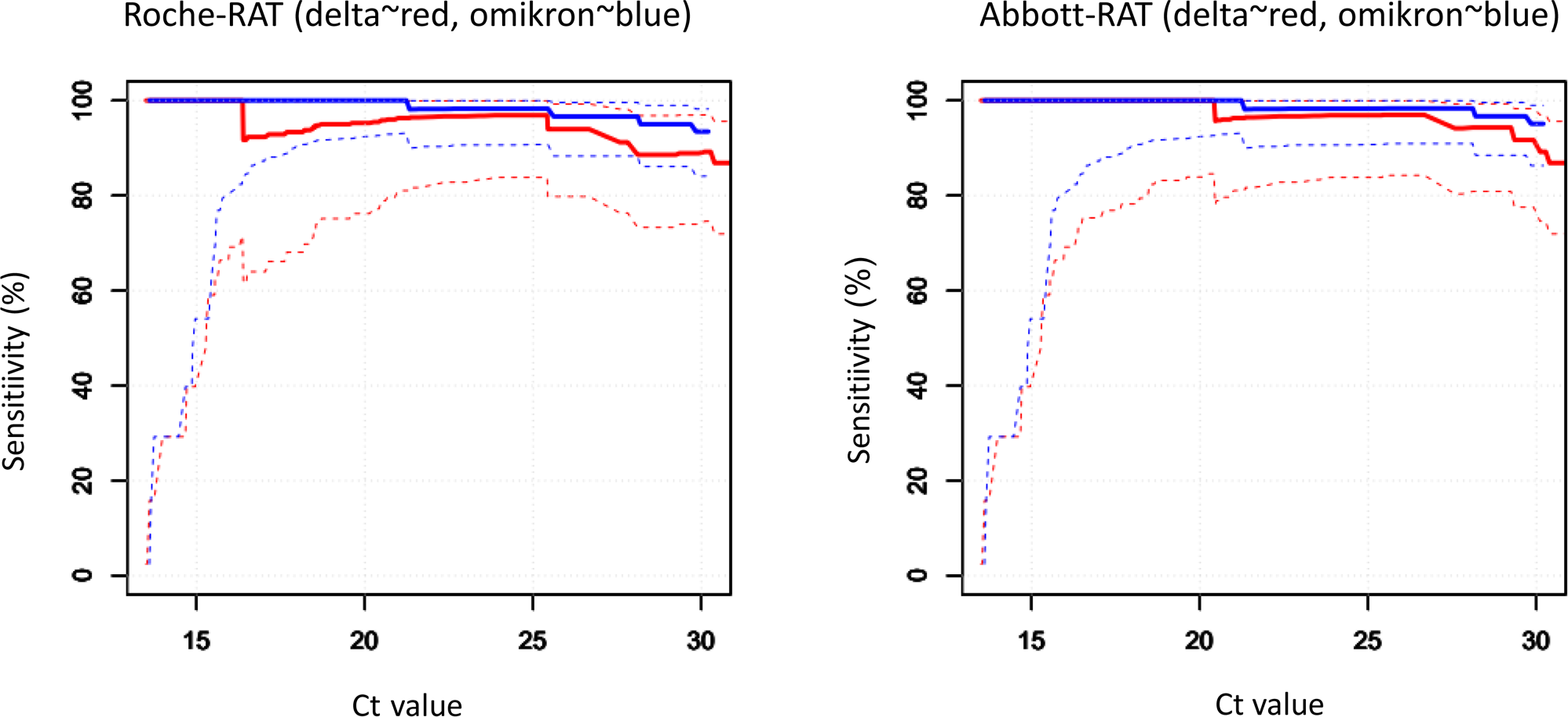
Relationships between sensitivities of RDTs vs. rRT-PCR cycle threshold (Ct) values. The solid lines indicate sensitivities, the dotted lines represent the upper, and the lower bounds the corresponding 95% confidence intervals. (A) left: Roche-RDT; right: Abbott-RDT. (B) Sensitivities according to SARS-CoV-2 genotypes. left: Roche-RDT; right: Abbott-RDT; *red*: Delta variant; *blue*: Omikron variant.

The Abbott-RAT and Roche-RAT did not show a significant difference in sensitivity (p=0.2091; Table 2). Due to higher power in the total study cohort (n=1487) the Roche-RAT had a significantly higher sensitivity than the Abbott-RAT (p=0.0093, Supplementary table 2). Among participants seeking testing due to a referral by their primary care physician, the sensitivities for the Abbot-RAT and Roche-RAT were 83.5% and 84.8%, for participants with a referral by the health department they were 54.6% and 57.6% and following a warning by their Corona-Warn-App the sensitivities were 48.8% for both tests (Table 2), respectively. In the participants excluded because they were tested to confirm a positive antigen test the sensitivities of the Abbott-RAT and the Roche-RAT were 93.0% and 94.5%.

Participants with at least one comorbidity showed lower sensitivities (49.1% and 52.8%, Abbott-RAT and Roche-RAT, respectively) than those with no comorbidities (71.9% and 72.6%%, Abbott-RAT and Roche-RAT, respectively; Table 2). This finding did not apply to individual comorbidities besides previous Covid-19. Participants with previous Covid-19 showed significantly lower sensitivities of only 26.7% for both RATs (OR 0.12 (95%CI: 0.05,0.3), p<0.0001). This finding is attributable to Ct values being markedly higher (Median 31.2) in patients with previous Covid-19 and not consistent anymore when adjusted for the Ct value (Table 3).

For participants without previous Covid-19, significantly higher sensitivities (72.8% and 74.7%, Abbott-RAT and Roche-RAT, respectively) were found in line with markedly lower Ct values (Median 19.2).

In symptomatic participants, the sensitivities were significantly higher (76.0% and 77.4%%, Abbott-RAT and Roche-RAT, respectively) than in asymptomatic participants (28.6% and 31.0%, Abbott-RAT and Roche-RAT, respectively). This finding is in line with Ct-values being lower in symptomatic patients than in asymptomatic patients (Ct Median 18.7 vs. 30.8, Table 2).

We further analyzed the diagnostic performance of RATs according to the vaccination status. The sensitivities of the RATs in non-vaccinated participants (0 or 1 vaccination) were 64.9% and 67.7% for Abbott-RAT and Roche-RAT, respectively. For participants with two vaccinations, the sensitivities were 59.4% and 60.9%. For participants with at least one booster vaccination, we find sensitivities equal to 70.6% for both RATs.

We also investigated whether the SARS-CoV-2 variants Delta and Omicron affected the sensitivity of the RATs. Both variants had similar sensitivities compared to the wild-type from the first wave of the Covag study. Compared to the alpha variant the alpha variant had significantly lower sensitivities than the wild-type, delta and omicron. (Figure 4). ^13^

**Figure 4:**
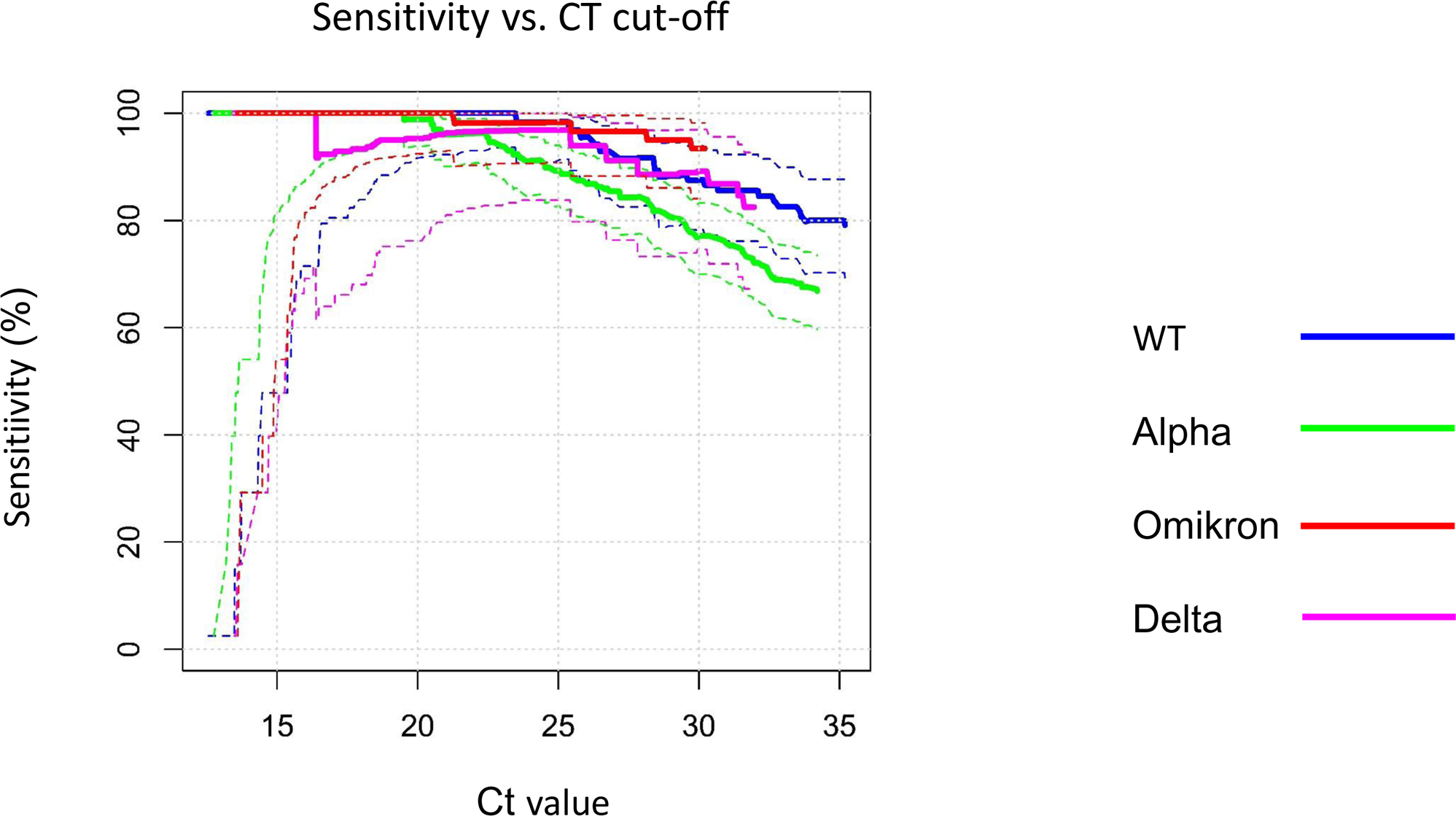
rRT-PCR cycle thresholds (Ct) values on rRT-PCR for SARS-Cov-2 RNA of different variants *versus* sensitivities of the Roche-RDT. The *solid lines* indicate sensitivities, the *dotted lines* represent the upper and the lower bounds the corresponding 95% confidence intervals. *magenta*: Delta; *red*: Omikron; *green*: Alpha; *blue*: WT.

To firmly establish independent predictors of sensitivity, we calculated ORs for having a positive RAT according to subgroups by multivariate logistic regression (Table 3). Covariables were age, sex, Ct value, reason for testing, presence or absence of any comorbidity and previous Covid-19, Covid-19 vaccination status, presence or absence of any clinical symptom, and the SARS-CoV-2 genotype. As expected, Ct values were strongly associated with the sensitivities of both tests. The sensitivities of the Abbott-RAT and Roche-RAT were lower in participants who sought testing due to a warning in the Corona Warn App.

When excluding the Ct value from the multivariate logistic regression symptomatic participants were detected with a significantly higher sensitivity than asymptomatic participants (Abbott-RAT: OR 4.35, p=0.0081; Roche-RAT: OR 3.46, p=0.0216). However, when adjusting for the Ct value this finding was not significant anymore (Table 3). The vaccination status was not associated with a change in sensitivity of the RATs.

As the Ct value is the strongest predictor for the sensitivity of the RATs, we calculated the sensitivity of the RATs separately for different Ct values. For a Ct value ≤ 25 the sensitivities were 95.2% and 96.0% for the Abbott-RAT and the Roche-RAT, respectively (Table 4). For a Ct value of 25-30 both RATs had a sensitivity of 18.8%. For a Ct value of 30-32, the sensitivities were 0.0% and 7.1% respectively, for Ct values ≥32 the sensitivities were 3.0% and 6.0% for Abbott-RAT and Roche-RAT, respectively.

Specificity. The specificity exceeded 99% overall and in mostly all participant strata (Table 2, Supplementary table 2).

PPV, NPV, and diagnostic performance. The rate of true negatives in our study cohort (n=888) was 697 of 700 (99.6%) and 698 of 700 (99.7%), the rate of false negatives was 65 of 188 (34.6%) and 62 of 188 (33.0%) for the Abbott-RAT and the Roche-RAT, respectively. The rate of true positives was 123 of 188 (65.4%) and 126 of 700 (67%). The rate for false positives was 3 of 700 (0.4%) and 2 of 700 (0.3%) for Abbott-RAT and Roche-RAT, respectively.

When also including the participants who already had a positive self-test beforehand (total of n=1487) the rate of false negatives decreased to 101 of 704 (14.4%) and 90 of 704 (12.8%) for the Abbott-RAT and the Roche-RAT, respectively. The rate of false positives was also overall very low with 4 of 783 (0.5%) and 2 of 783 (0.3%) for the Abbott-RAT and the Roche-RAT. Of the 591 participants who sought RT-PCR testing to confirm a positive self-test, 511 (86.5%) were confirmed positive by RT-PCR while 80 (13.5%) were tested negative by RT-PCR.

The SARS-CoV-2 prevalence in our study cohort was 78.8% (n=888). At this prevalence the PPV was at 97.6% and 98.4% for Abbott-RAT and Roche-RAT (n=888, Table 2). For symptomatic participants the PPV was higher (98.2% and 98.3%, Abbott-RAT and Roche-RAT, respectively) than for asymptomatic participants (92.3% and 96.3%, Abbott-RAT and Roche-RAT, respectively). The NPV was 91.5% and 91.8% for Abbott-RAT and Roche-RAT. The NPV was higher for asymptomatic (93.0% and 93.2%, Abbott-RAT and Roche-RAT, respectively) than for symptomatic participants (89.5% and 90.1%, Abbott-RAT and Roche-RAT, respectively).

Because patients with SARS-CoV-2 infections were enriched in our study population compared to the general population, we examined the PPVs and NPVs at assumed prevalence rates up to 0.05 (Figure 5). To compare the PPV and NPV of the RATs with standardized performance criteria, we also used the following hypothetical sensitivity and specificity levels (tiers 1-3) recommended by Kost15: tier 1, 90%, 95%; tier 2, 95%, 97.5%; and tier 3, 100%, ≥99% (Figure 5). At this prevalence rate, our results suggest a PPV and NPV of 88.9% and 98.2% for Abbott-RAT, and 92.5% and 98.3% for the Roche-RAT, the Roche-RAT displaying a higher PPV than the Abbott-RAT and both scoring higher than the hypothetical tiers 1 through 3, reflecting increases in NPV in the order of Abbott-RAT < Roche-RAT < tier 1 < tier 2 < tier 3. The NPVs ranged in the order of tier 3 > tier 2 > tier 1 > Roche-RAT > Abbott-RAT.

**Figure 5:**
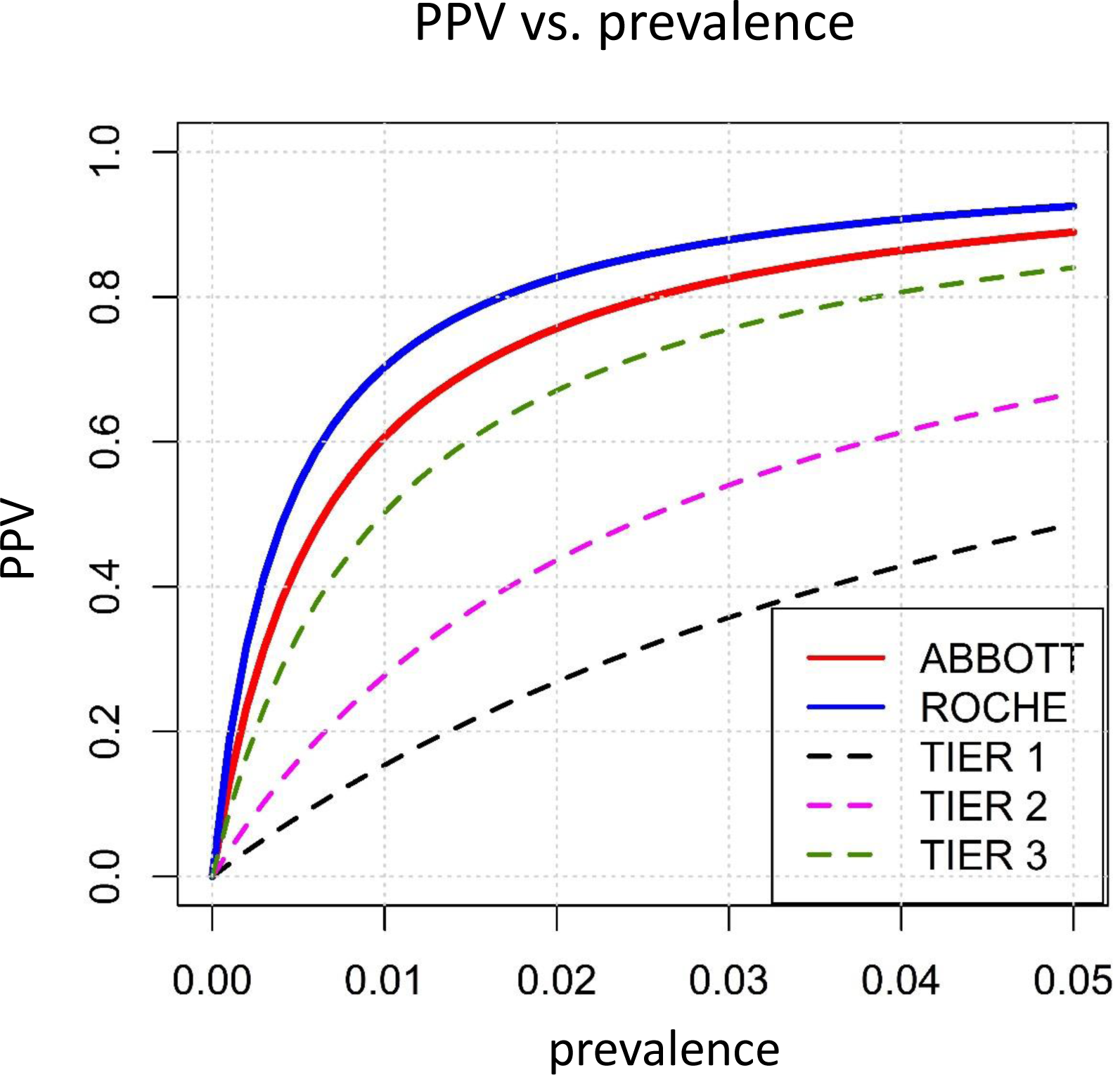
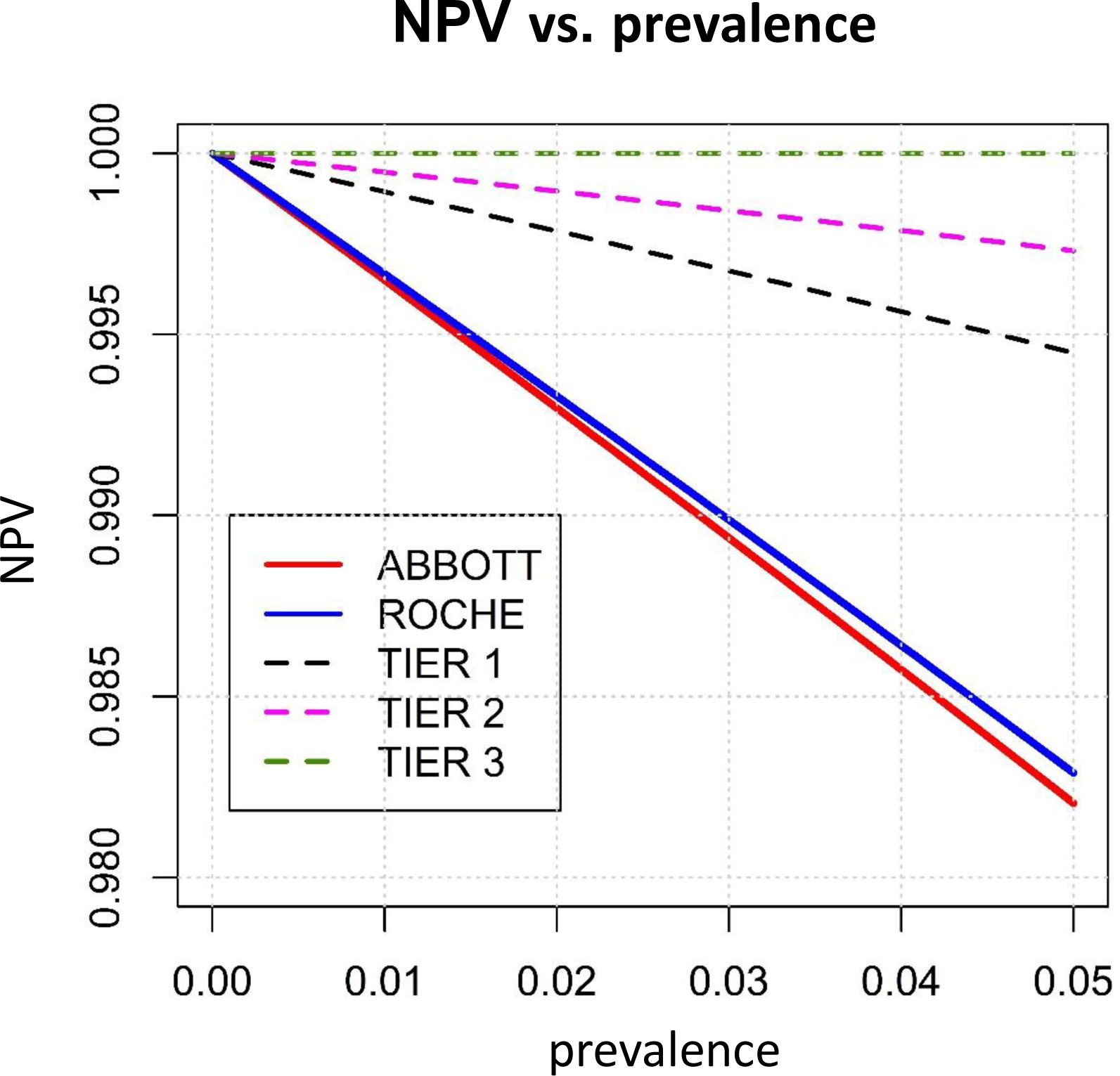
(A) Prevalence *versus* PPV for tier 1-3 (for hypothetical sensitivities and specifities (Kost et al.)) and both rapid tests. (B Prevalence *versus* NPV for tier 1-3 (for hypothetical sensitivities and specifities (Kost et al.)) and both rapid tests.

## 4. Discussion

This study is an extension of the COVAG study which is one of the largest prospective, real-world evaluations of RATs to date.^13^ We compared two of the most sensitive RATs provided by Abbott Diagnostics and Roche Diagnostics, especially in the light of newly emerged variants.9 We found that the sensitivities of RATs for asymptomatic patients was as low as 30%. We found that the Omicron and Delta variant were detected with not significantly different sensitivities compared to the wild-type at Ct values >25. 13

In contrast to the first wave of our study there was no significant difference in sensitivity between the Abbott-RAT and Roche-RAT.^13^ However, with an extended sample size (n=1487) after including participants seeking RT-PCR testing to confirm a positive antigen test, the Roche-RAT had a significantly higher sensitivity than the Abbott-RAT (p=0.0093; Supplementary table 2), attributable to the participants with an age ≤ median. This finding is in good agreement with the results of the first wave of our study.

The sensitivities were substantially higher among participants referred by their primary care physician (84-85 %, Table 2). As primary care physicians refer patients to RT-PCR testing based on their clinical presentation and history, the pretest probability is higher and patients with higher symptom burden sent for testing, also reflected by lower Ct values in these participants. This shows that the sensitivity of the RATs can be increased by considering the clinical background. The PPVs of RATs was overall very good (88-92%). Compared to the tiers recommend by Kost et. al the NPV occur to be lower than the recommend values of tier 1-3. However, due to the smallness of the discrepancies between the measured NPVs and the recommended NPV ranges (<2%) and the small number of false positives, the last statement made about the NPVs should be taken with caution.

### 4.1. Diagnostic performance of the RATs

The WHO formulated minimum performance requirements of ≥ 80% sensitivity and ≥ 97% specificity for RATs.^16^ The European Centre for Disease Prevention and Control (ECDC) agreed to the performance requirements set by the WHO.^17^ In our study both RATs did not meet the sensitivity performance requirements while meeting the specificity requirements (Abbott-RAT: sensitivity 65.4%, specificity 99.6%; Roche-RAT: sensitivity 67.0%, specificity 99.7%). Similar results were reported by a Cochrane Analysis which reported sensitivities of 56.7% (95% CI 44.3-68.3%) and 64.4% (95% CI 52.2-75.0%) for the Abbott-RAT and the Roche-RAT, respectively.^9^ In a large comparative <u>in vitro</u> evaluation of 122 RATs reported the Paul-Ehrlich-Institut (PEI), the overall sensitivity of the Abbott-RAT and the Roche-RAT were 64.0% and 46.0%, respectively.^18^ While the Abbott-RAT showed a comparable sensitivity of 65.4% in our study, the Roche-RAT yielded a better sensitivity of 67.0%. However, also in the study by the PEI both RATs failed to meet the sensitivity requirement set by the WHO. This is in large contrast to the sensitivities of 97.6% and 95.5%, respectively, reported by the providers Abbott and Roche. for samples with Ct values ≤ 30. ^19,20^

During our study comparable sensitivities (95.2% and 96% for Abbott-RAT and Roche-RAT, respectively) were reported only for Ct values ≤ 25. For Ct values of 25-30 the sensitivities were only 18.8% for both tests.

The RATś performance strongly relates to Ct values. The study by the Paul-Ehrlich-Institut showed sensitivities for the Abbott-RAT of 100% for Ct-values ≤ 25, 60.9% for Ct values between 25-30 and 0% for Ct values ≥ 30.^18^ The Roche-RAT in comparison yielded a sensitivity of 88.9% for Ct values ≤ 25, 30.4% for Ct values between 25-30 and also 0% for Ct values ≥30.^18^ Evidently thus, the performance of the RATs in our study is worse than in the <u>in vitro</u> study by the <u>Paul</u>-Ehrlich-Institut, suggesting that challengeable information will only be obtained under real world conditions. This notwithstanding the common denominator of the results fom Paul-Ehrlich-Institut and of ours is that the performance requirements are only met for samples with a Ct ≤ 25. Hence, patients with a high viral load are well detected while patients with a lower viral load are missed.^21^

An important clinical distinction is whether symptoms are present or not. The sensitivity of the RATs is markedly lower for asymptomatic than for symptomatic patients. With a sensitivity of around 30%, asymptomatic and infected patients were detected at very low sensitivity in our study. Symptomatic patients on the other hand are detected with a sensitivity of around 77%. A Cochrane analysis by Dinnes et al. reported similar results for symptomatic (Abbott-RAT: 74.8%; Roche-RAT: 78.8%) and higher results for asymptomatic (Abbott-RAT: 56.9%; Roche-RAT: 59.4%) patients compared to our study.^9^ Although slightly below the performance requirements of the WHO RATs may be considered useful in symptomatic patients while they are not in asymptomatic patients. These differences in sensitivity are clearly attributable to the lower Ct values of symptomatic patients. In Germany RATs have been used for screening of asymptomatic persons.^10^ Yet, in these patients RATs are clearly insufficient for screening.

The RNA viral load determined by RT-PCR is only a proxy for the infectiousness of patients as also non-infectious viral RNA is detected by RT-PCR. To reliably determine the infectiousness of a patient, viral growth can be examined in culture. In a study from the UK, contacts of SARS-CoV-2 infected patients were recruited, and RT-PCR and virus culture were performed daily. Additionally, a RAT different from the ones used in our study was performed in RT-PCR positive samples as well as in samples one day before and after a positive RT-PCR. The sensitivity of the RATs was higher for samples with positive viral cultures (79%) than for samples with only positive RT-PCR (47%). Positive viral cultures were detected for a median of 5 days (IQR 3-7 days) and the peak viral load determined by viral cultures and RT-PCR was at a median of 3 days after symptom onset (IQR 3-5/6 days). Interestingly the sensitivity of the RATs was lower before and during the peak viral load (sensitivity: 67%) than after the peak viral load (sensitivity: 92%).^22^ This shows that RATs have reduced sensitivity during the beginning of infection possibly leading to delayed diagnosis.^22^ In a study from Germany the Roche-RAT was compared to RT-PCR and viral culture. Although the Roche-RAT reached a sensitivity of only 42.8%, none of the samples with positive viral cultures was missed.^23^ Hence and accordance to the current study, RATs appear to have a low overall sensitivity, while highly infectious participants may reliably be detected.

The specificity of the RATs was overall very good and met the specificity requirements of the WHO and ECDC.^16,17,24^

### 4.2. Influence of the SARS-CoV-2 genotype on the diagnostic performance of RATs

During the first data collection period from February 1 to March 31, 2021, the dominant variants were the wild-type and the alpha variant. The sensitivities of the RATs for the alpha variant were significantly lower than for the wild-type also when adjusted for the Ct-value.^13^ In the current wave of our study ((October 20, 2021 to March 18, 2022), the prevailing variants were Delta followed by Omicron. Omicron was detected with a high sensitivity of 92-94%, while Delta was detected with a lower sensitivity of 80%. This difference can solely be explained by the lower Ct values of Omicron compared to Delta (Median 17.6, IQR 15.7-19.8 vs. Median 19.6, IQR 16.3-23). Consistently, in a multivariate logistic regression adjusted for the Ct values there was no significant difference between Omicron and Delta anymore. Also, when compared at set Ct values of ≤25, 25-30, ≥30 there was no significant difference in sensitivity for Delta and Omicron, respectively. While it has been argued that that Omicron produces a higher viral load leading to better detection by RATs in general, recent findings do not confirm this assumption.^25,26^ Another study from the USA also found that the sensitivities for Omicron compared to the Delta variant are not significantly different.^27^

We further examined the sensitivities for Omicron and Delta compared to the wild-type data coming from the first data collection period.

### 4.3. Influence of the Covid-19 vaccination and previous infection on the diagnostic performance of RATs

For patients with previous Covid-19 the sensitivities for Abbott-RAT and Roche-RAT were very low (26.7%). These low sensitivities are attributable to the high Ct values in these patients (Median 30, IQR 25-33). This is plausible because patients with a previous Covid-19 infection may have lower viral loads due to mucous IgA built in response to the previous infection.^29^ There was no significant difference in the sensitivities between vaccinated and unvaccinated participants (65-66% vs. 66-70%, Table 2)., perhaps since mucous IgA is formed to a lesser extent after vaccination.^30^ This could explain why the vaccination status does not seem to influence the sensitivity of RATs, while a previous Covid-19 infection could. Another explanation would be that after vaccination antibodies are formed only against the Spike protein whereas after a previous infection antibodies against the Spike protein and the Nucleocapsid protein are formed.^31^ As RATs detect the Nucleocapsid antigen Nucleocapsid antibodies could reduce available antigens for detection.

### 4.4 Limitations

Among the limitations of this study is that the reference method RT-PCR does not indicate the infectiousness of patients, because RT-PCR can also detect non-viable virus particles, also there is a certain correlation between the Ct value and infectivity.^32^

Furthermore, we performed RATs once only and not in series. Serial testing for SARS-CoV-2 with RATs may substantially increase their diagnostic performance.^33^

### 4.5 Conclusions

The diagnostic performance of RATs is highly associated with the viral load. The sensitivity of RATs is substantially higher in symptomatic than in asymptomatic patients and in patients referred by primary care physicians compared to other reasons for testing. Hence, RATs are significantly more useful in a clinical setting than for screening purposes. Our study does not suggest that the vaccination status influences the sensitivity of RATs.

## 6. Author Contributions

WM designed the study. CW, AS, HB, NL and H-JW collected the data. AD performed the statistical analysis. EW, AS, HB, NL surveyed the laboratory analyses. CW wrote the manuscript. All authors validated, reviewed, and edited the manuscript.

## 8. Acknowledgments

We thank the participants for joining in free of remuneration. We thank Katja Pöhl, CEO, SYNLAB MVZ Leinfelden-Echterdingen and Christoph Mahnke, CEO SYNLAB Holding Deutschland GmbH, who supported the study. We thank all study personel, especially Manuel Kraft, Ali Hussein, Alexander Ignatenko, Brigitte Schwandt, Orhan Bunjaku and Rosa Oberhauser for carrying out the RATs and documenting the participants at the Corona Test Center Cannstatter Wasen, Stuttgart.

## 9. Data Availability Statement

Data will be made available to researchers upon justified request and formal agreement to make sure that rules of good scientific practice are obeyed, and that credit is given to the people who have been in charge of the design and the organization of the study. Interested researchers are invited to address their request or proposal to WM. The authors confirm that they accessed and validated these data and that all other researchers can access the data in the same manner the authors did.

